# Monitoring of SARS-CoV-2 B.1.1.7 variant early-phase spreading in South-Moravian Region in the Czech Republic and evaluation of its pathogenicity

**DOI:** 10.1101/2021.05.24.21257365

**Authors:** Daniel Diabelko, Milada Dvorackova, Monika Dvorakova Heroldova, Giancarlo Forte, Ivan Cundrle, Filip Ruzicka, Jan Vrbsky

## Abstract

SARS-CoV-2 emerged in Wuhan, China, in December 2019. Starting in January 2020, over a period of several months, the initial virus (Wuhan-Hu-1/2019; Wu et al. 2020) diverged in a descendant strain carrying D614G amino acid mutation in spike protein. By summer 2020 this novel coronavirus (nCoV) became the most dominant form of the virus circulating worldwide and raised serious international concern. Currently (April 2021), there are 3598 subsequent PANGO branched lineages recognized that carry numerous mutations. To date, the most emerging lineages of SARS-CoV-2 worldwide include B.1.1.7 lineage with a frequency of 48% followed by several dozens of others with frequencies 7.5% or less, such as B.1.351, B.1.1.28, B.1.2, B.1.1.519, P.1, R.1, etc. (www.nextrain.org, Centers for Disease Control and Prevention; CDC 2020 www.cdc.gov).

In this study, we monitored the spreading of B.1.1.7 lineage from the early phase of its appearance until it became predominant in the South-Moravian region of the Czech Republic. We measured significantly associated clinical marker (Ct; cycle threshold) correlating with viral load in B.1.1.7 lineage. Interestingly, we found that the spreading of B.1.1.7 strain was associated with a shift in patients average age, as compared to the former predominant lineage. Finally, we calculated the impact of the B.1.1.7 lineage on hospitalization and case fatality of the patients on the intensive care unit in the central South-Moravian faculty hospital.

## Introduction

SARS-CoV-2 is an enveloped virus with non-segmented, positive-sense, single-stranded RNA. The genome consists of both structural and non-structural protein precursors, accessory protein precursors and 5′, 3′ untranslated regions. The ORFlab is a non-structural polyprotein precursor, which is translated into several mature proteins (nspl-16). These are essential for genome maintenance and replication of the virus. The structural protein precursors (Fig. 1) encode membrane (M), spike (S), envelope (E) and nucleocapsid (N) proteins. Spike protein consists from two domains, S1 and S2. Receptor binding domain (RBD) is located on the S1 domain and is responsible for Angiotensin-Converting Enzyme 2 (ACE2) binding. S2 domain facilitates viral fusion with the host cell (Yan et al. 2020, Xia et al. 2020). Therefore, spike protein is considered as the major factor of virulence and immunogenicity of SARS-CoV-2 virus.

**Figure 1.**
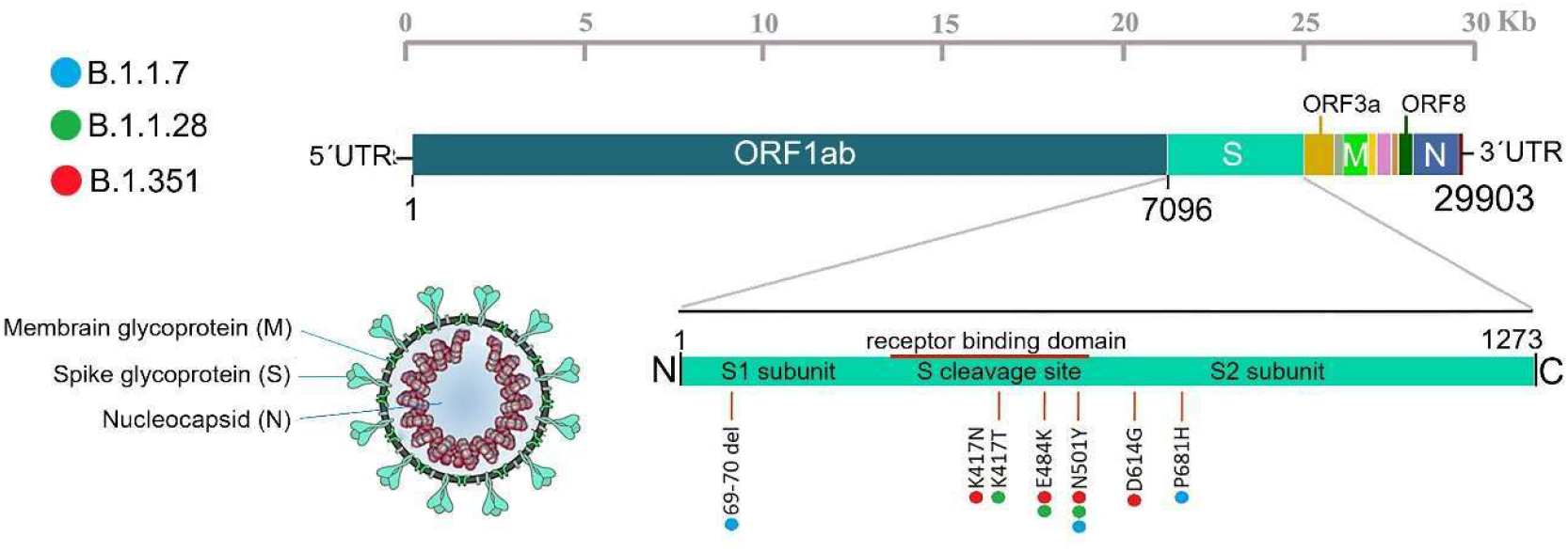
Schematic representation of SARS-CoV-2 genome structure and selected mutations. The viral surface proteins, envelope membrane glycoprotein (M) and spike glycoprotein (S), are embedded in a lipid bilayer, while the nucleocapsid (N) consists of single-stranded viral RNA (ss-RNA) associated with the nucleocapsid protein. The spike protein consists of S1 and S2 subunits and harbors cleavage sites for furin. Mutations defining actual PANGO lineages are color coded (adopted from Laamarti et al 2020).

Albeit numerous evidences were provided that novel SARS-CoV-2 mutations aroused in the past months, its mutation rate is estimated to be around 6-8 × 10-^**4**^ nucleotide substitutions per site per year (n/s/y) (van Dorp et al. 2020, Sanjuan 2012, Domingo-Calap et al. 2018, GitHub; https://github.com/nextstrain/ncov). This mutation rate is twice lower than influenza A, which ranges from 1.43×10−3 n/s/y (H3N8) to 11.62 × 10-3 n/s/y (H7N7) (Rejmanek et al. 2015), or SARS-CoV, which is credited of 0.8-2.38 × 10-3 n/s/y (Zhao et al. 2004). Overall, mutation rate estimates of SARS-CoV-2 are rather below average compared to the other positive-sense RNA viruses.

The impact of individual mutations on the virus behavior is poorly understood. Of the highest interest are the mutations in S-gene that were reported to result in conformation changes in the Receptor-Binding Domain (RBD) within the spike protein and increased virulence such as increased infectivity and higher viral load in infected patients (Yurkovetskiy et al. 2020, Korber et al. 2020, Volz et al. 2021, and many other authors). One of the significant symptoms attributed to novel mutations is an impact on chemosensory dysfunction (Butowt et al. 2020, van Bartheld et al. 2020). The information whether and if, to what extent newly accumulated mutations lead to a decrease in immunogenic capacity or increase in mortality is considered of great relevance, especially as part of the COVID-19 vaccines design process.

Lineage B.1.351 was originally detected in South Africa, where it shortly became the predominant variant (Tegally et al. 2020). Among others, it carries NS0lY, K417N, E484K, and D614G defining amino-acid substitutions (CDC 2020). This lineage displays increased transmissibility and can affect antibody-driven neutralization with the potential to reduce the efficacy of some of the vaccines available on the market (Weisblum et al. 2020, ECDC 2021, Jangra et al. 2021, Liu et al. 2021). Until mid-April 2021, its spreading was detected by sequencing in 71 additional Countries in nearly ten thousand sequenced samples (PANGOLIN, April 2021).

Lineage B.1.1.28 was first detected in Brazil. Its defining mutations include N501Y, E484K, and K417T (Tang et al. 2021). It was shown to have increased transmissibility and to possibly reduce vaccine efficacy (Greaney et al. 2021, Jangra et al. 2021, Liu et al. 2021). This variant was detected in 37 countries (mid-April, 2021) in 3770 sequenced samples worldwide (PANGOLIN, April 2021).

Lineage B.1.1.7, firstly detected in the United Kingdom, seems to be the fastest spreading of these lineages mentioned (Openshaw et al. 2020). Until April the 20th 2021, it was detected by sequencing in 113 countries worldwide, reaching over three hundred thousand sequenced samples (PANGOLIN, April 2021). The most notable mutations in this lineage include N501V, del69/70, and P681H (Chand et al. 2020). Ascribed novel features of this variant include increased efficiency and rapid transmission caused by stronger affinity to the ACE2 receptor, resulting in a further increase of its infectivity by up to 75% (Leung et al. 2021, Kemp et al. 2021). This significantly strengthens the pandemic situation caused by SARS-CoV-2. Furthermore, lineage B.1.1.7 was associated with an increased risk of death (Horby et al. 2021, Davies et al. 2021).

Despite the enormous effort of thousands of researchers in the covid field, still many questions remain to be answered in order to control COVID-19 pandemics. In the following paragraphs, we describe the capacity of B.1.1.7 lineage to spread and overgrow other variants of SARS-CoV-19 on the model of the South Moravian Region of the Czech Republic, with a particular focus on a description of selected clinical and diagnostic markers.

## Results

### Initial spreading of B.1.1.7 lineage in South-Moravian region of the Czech Republic

Lineage B.1.1.7 was first detected in the Czech Republic in first week of January 2021 (Jirincova, Paces 2021) after its initial spread from the United Kingdom and other north-west neighboring countries (Figure 1a) (GISAID, Nextstrain). Incidence of the B.1.1.7 variant differs among districts in the Czech Republic (Nachod 45%, Trutnov 60%, Prague <10%) (Jirincova, Paces 2021). First detection of B.1.1.7 in our laboratory happened on the 22^nd^ of January 2021. Starting from the 1st week of February, we detected massive increase of the SARS-Cov-2 cases with B.1.1.7 lineage. As documented on period of 10 days only (from February 3^rd^ to 13^th^ 2021), we measured a 40 % increase in B.1.1.7 positive cases. By the end of February and 1st week of March, this lineage almost exclusively replaced the original lineages (predominantly B.1.258 - 52%, followed by minor incidences of B.1, B.1.1, B.1.221, C.35, B.1.527, B.1.1.277, B.1.177, B.1.160, B.1.258; NRL, Nextstrain) reaching nearly 97% **(Figure 2b)**. In this period, total SARS-CoV-2 positivity in the Czech Republic reached 1572 per hundred thousand people (Komenda et al. 2021). We concluded that in this relatively short period we witnessed the community spreading of B.1.1.7 lineage in South-Moravian region in the Czech Republic, as calculated from 1539 samples positive for SARS-CoV-2, from which 1003 were identified as B.1.1.7 lineage.

**Figure 2.**
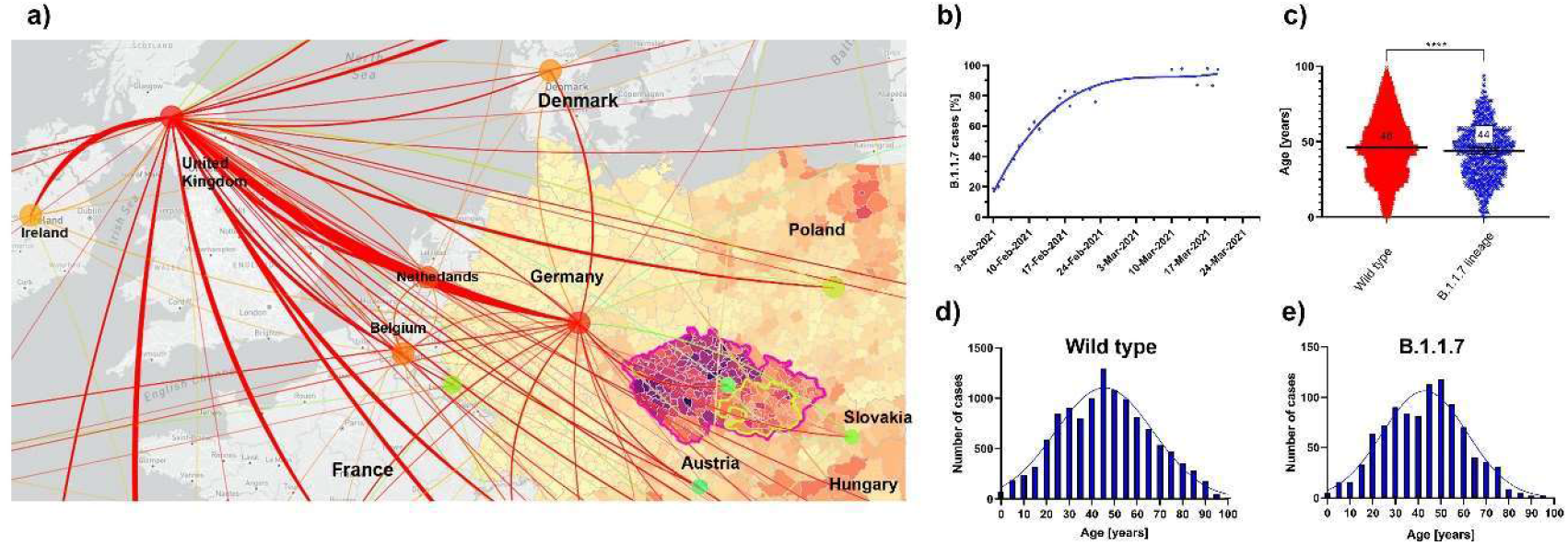
The spreading of B.1.1.7 lineage in the Czech Republic and change in age distribution of B.1.1.7 positive patients. a) Scheme of the B.1.1.7 spreading in western and central Europe. Color heatmap in Germany, Poland, Austria, Czech Republic, Slovakia and Hungary represents the number of new weekly cases per 100.000 people as of March 4^th^ 2021. Modified from Mahdalova K. (https://www.seznamzpravv.cz based on data from Robert Koch Institute, OGD Österreich, ÚZIS, MC Polska) and GISA/0 transmission map of B.1.1.7. b) Graph of B.1.1.7 identification rate from February 3^rd^ to March 26^th^ (n=1539 samples tested, 1003 were identified as B.1.1.7 lineage). c) Scatter dot plot, Median with 95% Cl representation of age distribution of B.1.1.7 and Wild type patients. Mann-Whitney test, p < 0.0001, n = 11699 for wild-type, 980 for B.1.1.7 samples. Median values are indicated. d, e) Histogram representing the age distribution of infected patients with B.1.1.7 and wild type lineages; n = 980 for B.1.1.7 and n = 11699 for wild type patients.

We analyzed the age distribution in cohort of 11699 cases of major lineages referred as “wild type” and 980 cases of B.1.1.7 lineage. We found a significant reduction of 3.7 years and median difference of 2.0 years **(Figure 2c)**. The age distribution of patients in both groups is shown in **Figure 2d** and **e**.

### B.1.1.7 lineage is correlated with increased viral load

To test whether the viral load in patients with the SARS-CoV-2 differs between B.1.1.7 and wild type lineages, we compared 1056 samples from which 863 were identified as B.1.1.7 lineage, while 193 as wild-type. We measured cycle threshold (Ct) and calculated a median difference of 8.45 cycles (Mann-Whitney U test, p<0.0001; figure 2a). With theoretical PCR amplification efficacy (2^Ct^), this accounts for approximately 350 times higher viral load in the B.1.1.7 lineage. The comparison of the age distribution showed that B.1.1.7 patients are nearly 5.4 years younger than wild type lineage patients (mean age 44.5 vs. 49.9 years; figure 3b). We did not detect significant differences of Ct values in certain age groups (figure 3c). These results suggest a significant impact of B.1.1.7 lineage on increased SARS.CoV2 virulence.

**Figure 3.**
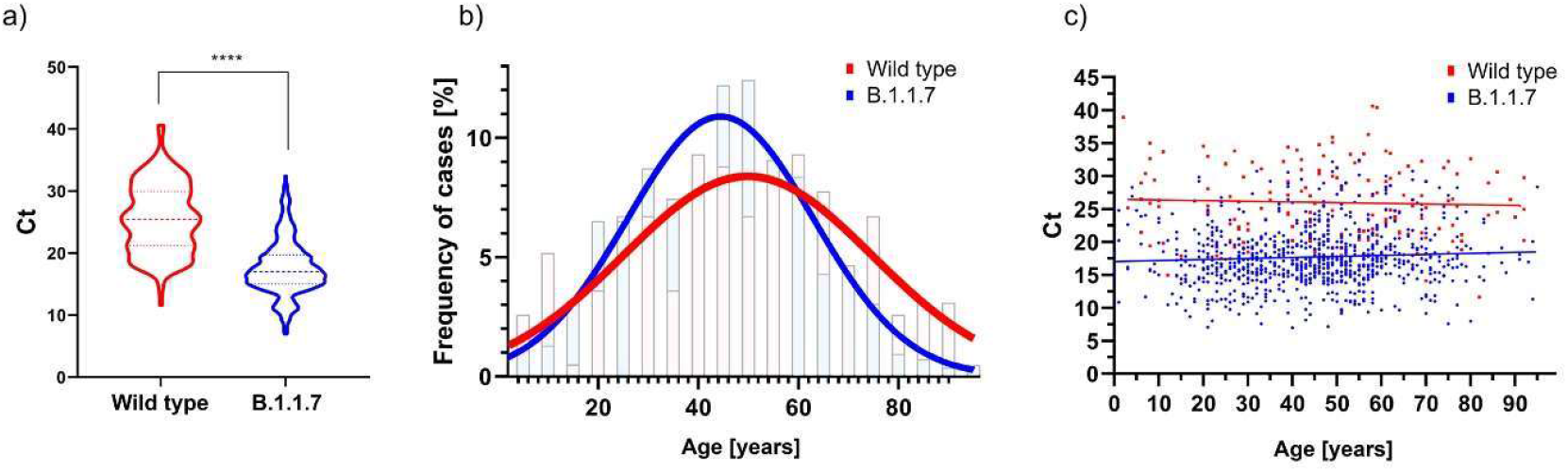
Comparison of Ct distribution of B.1.1.7 lineage and wild type. a) Violin plot of Ct distribution in wild type and B.1.1.7 lineages; the data are shown as median (193 for wild type, 863 for B.1.1.7) +/- quartiles (p < 0.0001). b) Histogram indicating the age distribution of wild type and B.1.1.7 positive patients overlayed with Gaussian curve from viral load analysis. c) Correlation graph representing the age and cycle threshold within B.1.1.7 and wild type group from Ct analysis. The overlayed linear curve represents a guide for the eyes.

### Evaluation of B.1.1.7 lineage effects on the number of ICU hospitalized patients and case fatality rate

In order to see the effect of COVID-19 B.1.1.7 lineage on the hospitalization and case fatality, we performed clinical monitoring of intensive care unit (ICU) hospitalized patients and compared the results to wild type lineage. We evaluated the most severe patients on ICU unit with SARS-CoV-2 interstitial pneumonia as primary diagnosis with comparable state at the start of hospitalization.

Age comparison of both cohorts of indicated age interval 25 to 82 years unveiled significantly younger patients hospitalized with B.1.1.7 lineage (median difference 58 vs 64 years of age, p < 0.05; **Figure 4a)**. Patients in both groups were selected based on breath support used (OTI, UPV, ECMO, HFNO, oxygen mask) and Oxygenation index (OI; calculated from FiO_2_, and PaO_2_; see methods for details), which was similarly represented in both groups **(Figure 4b)**. In the tested period of forty days, we counted 43 hospitalized patients with wild type lineages and 41 hospitalized with B.1.1.7 lineage (see methods for details of tested periods). The resulting number of hospitalized patients with severe symptoms caused by SARS-CoV-2 in our ICU was statistically indistinguishable between wild type and B.1.1.7 lineages (51 patients with B.1.1.7 versus 60 patients with control lineages).

**Figure 4.**
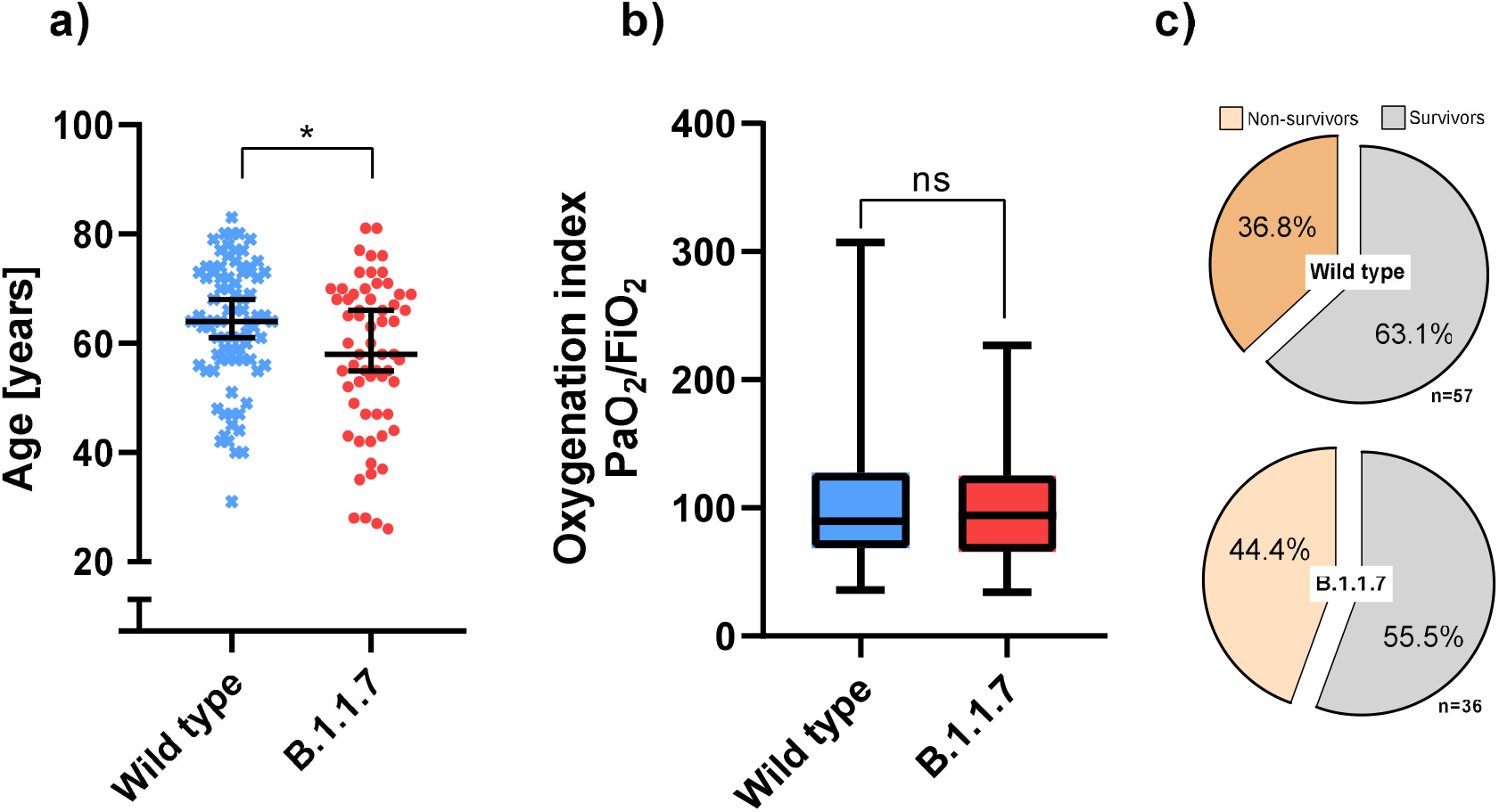
a) Scatter plot of age distribution of B.1.1.7 and wild type group. Median is indicated by the line and whiskers show 95% Cl interval. Mann-Whitney test, p = 0.01, median difference 6 years. b) Box-plot of oxygenation index of ICU hospitalized patients with both, B.1.1.7 a wild type lineage. Whiskers are showing min to max range. Median difference 1.95, Mann-Whitney test, p = 0.53. c) Case fatality rate in B.1.1.7 and wild type group. Percentage of survivors and non-survivors are indicated.

To evaluate the case fatality rate of B.1.1.7 lineage we counted odds ratio of death and released patients in ICU unit. In order to eliminate the effect of initiated vaccination of elderly patients (70+) these were excluded from the analysis. We counted odds ratio (OR= 1.37; Cl: 0.5808-3.215, p = 0.52) of deaths and survived patients for wild type (21/36) and B.1.1.7 lineage (16/20). We found no statistically significant difference among groups tested with trend of 7.6 % increase in B.1.1.7 lineage **(Figure 4c)**.

## Discussion

Due to the genetic variants of SARS-CoV-2, some of which were shown to modify viral characteristics, it is crucial to follow and evaluate their spread among and within countries. In the present study, we have shown how quickly the B.1.1.7 lineage became the most dominant variant on the model of South Moravian region of the Czech Republic. In a period of three weeks, the wild type variants were almost completely overcome by B.1.1.7 lineage, which was eventually detected in 97% of SARS-CoV-2 positive samples tested at our premises. In our laboratory, we test three to eleven hundreds of samples per day, mainly from the patients indicated for SARS-CoV-2 test by general practitioners and Public Health Office, as well as from patients hospitalized in St. Anne university hospital.

Based on the predicted changes induced by the mutations in the shape of the coronavirus spike protein (Singh et al. 2021), which the virus uses to attach and fuse with the host cells, we predicted B.1.1.7 lineage infection would be associated with higher viral load. Indeed, our calculations showed a statistically significant increase of more than three hundred times more viral copies. Our findings correlate with those reported by Kidd et al. 2021 in a different cohort of patients. By further analysis of B.1.1.7 patients, we found a median reduction of two years of age in infected people. This suggests B.1.1.7 lineage might be associated with higher infectivity, most likely caused by easier facilitation of virus/cell fusion resulting in higher number of infected cells as suggested by Leung et al. 2021, Kemp et al. 2021.

To better evaluate how B.1.1.7 lineage impacts on the number of patients hospitalized with COVID-19, we selected patients from intensive care unit primarily diagnosed with SARS-CoV-2 interstitial pneumonia. This strategy was adopted to better differentiate the effect caused by COVID-19 from other co-morbidities. Despite of the biological advantages and enhanced transmission discussed above, we confirmed no statistical significance in the number of hospitalized patients. No effect was found even when we excluded elderly patients (70+) in order to eliminate the effect of the vaccination campaign.

Both groups showed similar oxygenation index, which suggests comparable initial state of disease. The resulting odds ratio of 1.37 might suggests an effect of B.1.1.7 lineage on case fatality, although no statistical significance was confirmed. However, the data of Horby et al. 2021 and Davies et al. 2021 indicate increased risk of death correlated with B.1.1.7 lineage. We speculate that a higher number of patients sorted in age groups needs to be tested in order to prove the effect. Additionally, when we inspected other co-morbidities in the patients (i.e., hypertension, endocrine, nutritional and metabolic diseases, urinary tract infection and neoplasms), we could not show any characteristics suggesting overall differences among the groups tested.

In order to prove statistical significance of indicated trend of younger patients hospitalized with severe symptoms caused by B.1.1.7 lineage and higher case fatality rate of younger patients from B.1.1.7 group, greater number of patients would need to be evaluated.

The B.1.1.7 lineage was firstly detected in the first weeks of January 2021. From calculation of the basic reproduction number Ro for the Czech Republic we can see increase of Ro from 15^th^ January 2021 to 27^th^ of February 2021. This rise occurred despite of various epidemiological regulations and it is very likely to be caused by the spread of the B.1.1.7 lineage. By the end of February, the rising number of positive cases as well as the appearance of other variants such as B.1.1.28, B.1.351 and the massive spread of B.1.1.7 lineage, resulted in strengthening of epidemiological regulations in the Czech Republic which possibly led to the decrease of Ro later in March. This also correlates with our findings, detecting B.1.1.7 lineage on January 22^nd^ 2021 and the following rapid spread of the B.1.1.7 lineage in the first weeks of February (see Figure 5).

**Figure 5.**
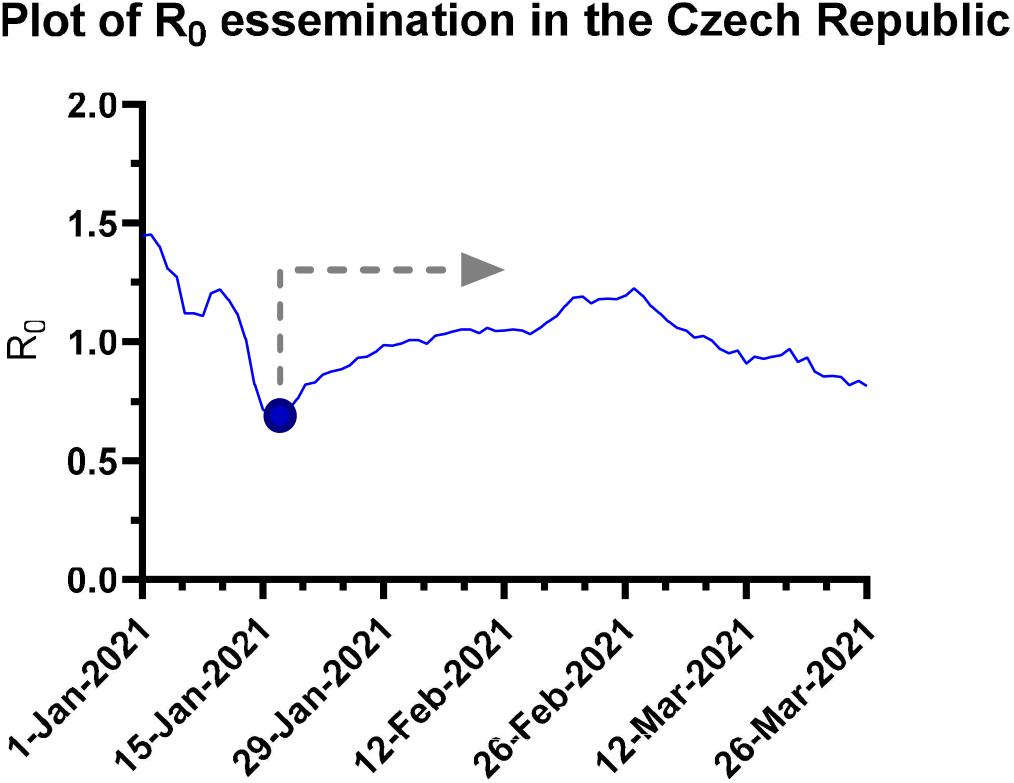
Spreading of B.1.1.7 lineage results in R number growth. In parallel with increase of B.1.1.7 lineage since January 18^th^ 2021 (marked by arrow) we could monitor the growth of basic reproduction number (basic reproduction number, calculated by Komenda et al. 2021).

We believe that results provided in this study inter-connecting molecular parameters of the B.1.1.7 lineage with clinical data from patients with known anamnesis provide valuable details which is frequently missing in global statistical studies.

## Material and methods

### Age selection of patients

For the age-distribution study, we analyzed hospitalized patients and people tested in COVID diagnostic units, thus includes patients with symptoms indicated by a general practitioner and self-payers. During the period from 3/2/2021 to 26/3/2021, we compared 1539 SARS-CoV-2 positive samples from which 1003 samples were identified as B.1.1.7 lineage. The age of 23 patients was not stated in documentation thus they were excluded from the statistic. The selection of the samples was done randomly. The B.1.1.7 age distribution was compared with the age distribution of all positive SARS-CoV-2 (wild type) patients from 20.3.2020 to 31.12.2020. (result significance unpaired Mann-Whitney test p < 0.0001, mean difference 3.676, median difference 2). This period was chosen to prevent any interference with B.1.1.7 lineage.

### RNA extraction and RT-PCR

RNA extraction was performed with Viral Nucleic Acid Extraction Kit (Zybia, Inc.) according to the manufacturer’s instructions. For RT-PCR analysis we used 5 µL of isolated RNA mixed with 15 µL of master mix Bio-Speedy® SARS-CoV-2 N501Y Mutation Detection Kit (Bioexen LTD) detecting both, N501Y and HV69-70del positive cases. For distinguishing other SARS-CoV-2 lineages we used Power Chek™ SARS-CoV-2 S-gene Mutation Detection Kit R6907Q (KOG-R6907Q-LS, Kogenebiotech), multiplex system recognizing N501Y, E484K and P681H mutations found in B.1.1.7, B.1 .1.28, and B.1.351 lineages respectively. To verify the authenticity of the samples, we performed NFS sequencing of 17 selected samples in collaboration with NRL, CEITEC, and Biogen.

### Viral load calculation

For viral load calculation, 1056 SARS-CoV-2 positive samples were chosen randomly. We identified 193 samples as wild type and 863 as B.1.1.7 lineage by RT-PCR with mutation specific primer pairs and under selective annealing conditions based on manufacturers protocols. Cycle threshold (Ct) values of all samples were calculated with the same optimized fluorescence threshold value (log RFU FAM 10-2) which refers to the viral load and which is designed to conserved genomic Orfl.2 region, where both B.1.1.7 and control samples share 100% homology. We further validated Ct values of wild type and B.1.1.7 lineage with the unpaired Mann-Whitney test.

### ICU Hospitalization and case fatality evaluation

Selection of ICU patients with B.1.1.7 and wild type lineages was done by the comparable initial state of pneumonia, breathing support, and primary diagnosis which was SARS-CoV-2 interstitial pneumonia. Oxygenation index calculated from the fraction of inspired oxygen (FiO2) and partial pressure of oxygen (PaO2) by the following ratio of PaO2/FiO2. PaO2 was obtained as the worst value in the first 24 hours of hospitalization in the ICU. We considered oxygenation ratio as the initial severity state of SARS-CoV-2 interstitial pneumonia. Selection of samples was also made based on types of breath support (orotracheal intubation, mechanical ventilation of the lung, extracorporeal membrane oxygenation, high-flow nasal cannula, breathing mask) that was similarly indicated in both groups.

SARS-CoV-2 vaccination on case fatality rate, we eliminated people older than 70 years. Statistical significance of odds ratio was evaluated by Fisher’s exact test and Baptista-Pike method for Cls of 95%.

To evaluate B.1.1.7 lineage effect on hospitalization, we chose similar periods of 40 days characterized with the predominant incidence of either wild type or B.1.1.7 lineage, respectively (Table 2). In both periods, the total number of SARS-CoV-19 positive cases confirmed by our laboratory was comparable (5582 for wild type or 5431 patients with B.1.1.7 lineage respectively). These numbers are in agreement with the statistics from the Ministry of Health of the Czech Republic (82 355 active cases on October 20th, 2020, and 86 805 active cases on February 20th, 2021; Komenda et al. 2021).

**Table 1.**

| **Case fatality rate** | Wild type | B.1.1.7 |
| --- | --- | --- |
| Test period | 92 days (1/10/2020 - 31.12.2020) | 44 days (25/2/2021-9/4/2021) |
| SARS-CoV-2 positive/total tested | 11270/40295 | 5862/19297 |
| ICU hospitalized | 57 | 36 |
| Deaths | 21 | 16 |

**Table 2.**

| **Hospitalization rate** | Wild type | B.1.1.7 |
| --- | --- | --- |
| Test period | 40 days (25/10/2020-3/12/2020) | 40 days (25/2/2021-5/4/2021) |
| SARS-CoV-2 positive/total tested | 5582/20378 | 5431/17804 |
| ICU hospitalized | 43 | 41 |

## Data Availability

The data that support the findings of this study are available on request from the corresponding author.

